# Brief Physical Activity Selectively Modulates the Performance of Serial Subtract 7 in Young Adults – A Wearable Sensor-based, Randomized, Control Study

**DOI:** 10.1101/2024.03.20.24302631

**Authors:** Xin Ran Chu, Tanmoy Newaz, Elbert Tom, Allison Yang, Taylor Chomiak, Bin Hu

**Affiliations:** Canadian Open Digital Health (OpenDH) program, University of Calgary, Calgary, Alberta, Canada T2N 4N1; Faculty of Nursing, University of Calgary, Calgary, AB T2N 4N1, Canada; Department of Family Medicine, Department of Community Health Sciences, Cumming School of Medicine, University of Calgary, Calgary, AB T2N 4N1, Canada; Faculty of Kinesiology, University of Calgary, Calgary, AB T2N 4N1, Canada; Department of Psychology, Faculty of Arts, University of Calgary, Calgary, AB T2N 4N1, Canada; Division of Translational Neuroscience, Department of Clinical Neurosciences, Hotchkiss Brain Institute, University of Calgary, Calgary, AB T2N 4N1, Canada

## Abstract

**OBJECTIVE:** This study explores the effects of physical activities on cognitive performance in healthy subjects, specifically evaluating Serial Subtract 7 Test (SST) performance during a cognitive-stepping dual task influenced by the 6-Minute Walking Test (6MWT) with and without music.

**METHODS:** A controlled experiment was conducted using the Ambulosono device to standardize walking exercises. 54 high school students participated, undergoing the 6MWT in different scenarios: Verbal 6-Minute Walking Test (6MWT) or Music-Guided Walking (MU). Final data from 43 students was used in the analysis. The SST measured cognitive changes in both single-task and dual-task conditions.

**RESULTS:** The 6MWT significantly enhanced cognitive performance in both single and dual-task conditions. However, the addition of music did not show a substantial improvement in cognitive performance. The findings indicated the positive impact of 6MWT on cognitive abilities, irrespective of musical accompaniment.

**CONCLUSIONS:** This research contributes to the understanding of how physical exercises can modulate cognitive functions in healthy individuals. It highlights the potential of 6MWT in enhancing cognitive performance, suggesting further exploration into the role of physical activity in cognitive health.

## Introduction

Cognitive faculties such as verbalization, working memory, comprehension, problem-solving, and decision-making are subject to age-related decline thereby compromising functional autonomy (Curzel et al., 2013; Murman, 2015; Roy, 2013). Numerous studies have indicated that cognitive functions can be substantially improved via non-pharmaceutical interventions such as aerobic exercises (Mandolesi et al., 2018; Wang et al., 2022). Physical exercise can augment cerebral health by fostering brain neuroplasticity through enhancing cerebral blood flow and overall well-being (Chang & Etnier, 2009; Colcombe et al., 2006; Erickson et al., 2011; Mandolesi et al., 2018). For example, a study by Mualem et al. (2018) showed that ten minutes of walking at a preferred speed can significantly improve memory and performance in critical feature-detection tasks in students of all age groups, suggesting that even brief exercise episodes can influence cognitive and academic performance (Mualem et al., 2018). Moreover, cardiorespiratory fitness can positively influence cognitive function in children as young as 4-6 years old (Keyes et al., 2021). A meta-analysis by Xu et al. (2023) further corroborated the cognitive benefits of physical exercise across the aging spectrum, irrespective of cognitive status, and advocated for adherence to current exercise guidelines.

Quantitative evaluation of exercise-cognition interactions and outcomes often faces a challenging technical issue, namely how to minimize the outcome variabilities caused by human delivery of instruction sets, the lack of standardization and cognitive testing protocols, as well as variety in exercise prescriptions (Herold et al, 2021, Montero-Odasso et al., 2023). Ambulosono is a smartphone-based wearable device and application system initially developed for evaluating cognitive-motor interactions and exercise intervention in Parkinson’s disease (Chomiak et al., 2019). The system allows standardization, automation, and non-human delivery of instruction sets, thereby reducing the influences arising from the testing environment (e.g. space and walkway limitations) as well as other technical issues that might skew the findings of traditional testing (Hu, 2019).

The present study aims to examine the utility of the Ambulosono system through the evaluation of how exercise can influence cognitive performance in healthy subjects. To this end, we constructed and implemented a set of testing and intervention protocols based on the Serial Subtract 7 Test (SST) (Haymen, 1942) and a dual task test (Ahman et al., 2020), in response to two different conditions of 6-minute walk exercises.

The SST is a standard evaluation of concentration, attention, and short-term memory (Chomiak et al., 2015; Haymen, 1942; Karzmark, 2000). It requires subjects to subtract 7 from 100 within a specified period, which consumes substantial attention and memory resources due to the demand for concurrent mathematical deduction and abstract reasoning while performing repeated subtraction (Chomiak et al., 2015; Haymen, 1942; Karzmark, 2000). As such, SST is often employed as a psychometric testing tool and diagnostic measure in both healthy subjects and patients (Graham et al., 2018; Srygley et al., 2009; Voelcker-Rehage et al., 2016). Dual-task performance is also an important cognitive measure that tests an individual’s capability to perform two tasks simultaneously (Ahman et al., 2020). Dual-tasking can lead to a decline in one or both task performances due to limited human attention resources and/or cognitive reserve (Ahman et al., 2020; Chomiak et al., 2015). It is generally believed that the behavioral manifestations captured in the presence of dual-task interferences reflect not only an individual’s ability to allocate attention resources but the degree of the attention dependence of the underlying mono-task during a dual-task test (DDT) (Al-Yahya et al., 2011; Pummer & Eskes, 2015). Variability in DTT performance, known as dual-task interference (Al-Yahya, 2011; Plummer & Eskes, 2015), is particularly evident in populations with neurodegenerative conditions such as Alzheimer’s, Parkinson’s, or those with post-stroke neurological deficits (Ahman et al., 2019; Yang & Pang, 2016). Clinically, DTT can be used to identify gait irregularities, fall risks, and cognitive impairments, which can guide therapeutic decisions (Yogev et al., 2007). Our study constructed a cognitive-motor dual-task test (DTT) by combining SST with stepping in place.

Although considerable information is available on how to use SST to construct a sensitive cognitive-motor DDT, relatively few studies examine whether such a DDT can be modulated by non-pharmaceutical interventions, such as exercise and music (Chomiak et al., 2015). This is despite the well-established theory that physical exercise can augment brain health by fostering neuroplasticity through enhancing cerebral blood flow and overall well-being (Chang & Etnier, 2009; Mandolesi et al., 2018).

In studying the effect of exercise interventions, defining the exercise type and intensity is essential. In our study, aerobic exercise was implemented via a 6-Minute Walking Test (6MWT). The 6MWT is a cost-effective, safe, and simple exercise evaluation that can elicit up to 80% of maximal heart rate and qualifies as a moderate to high-intensity exercise regimen (Sperandio et al., 2015; Wu et al., 2003). The Ambulosono device digitizes 6MWT, thereby controlling the exercise intensity using verbal speed instructions and capturing walk data via its wearable inertial sensors.

Apart from exercise, music listening is often cited as another form of lifestyle intervention for promoting cognitive, emotional, and psychosocial well-being (Särkämö, 2018). Cognitive domains such as working memory, processing speed, mood, and attentional control are thought to be positively modulated by musical stimuli although recent studies have refuted the previous claims such as the Morzat effect on attention (Mammarella et al., 2007; Steward et al., 2006; Thompson et al., 2005). Research also shows that listening to music during exercise can increase motivation and effort, while tempo-paced synchronous music can reduce perceived exertion, increase endurance, and increase the intensity and duration of exercise (Alter et al., 2015; Ballman, 2021; Hu & Chomiak, 2019). To control for the influence of cognitive arousal, a music walk group was also included in our study, in which the 6MWT instructions were replaced with music listening, with uniform song selection. We used a pre- and post-intervention design to investigate the changes in SST performance in healthy subjects under single or dual-task conditions following verbal or music walking.

Our pilot data indicates that SST and DTT can be used as sensitive and quantitative indicators for evaluating short-term influences of exercise on cognitive function. Furthermore, emotional interference of music listening during exercise can “mask” the benefits conferred by exercise on SST performance.

## Methods

### Experimental Design

Our study was conducted at a single site and received ethical clearance and informed participant consent via the Canadian Medical Hall of Fame, which hosted a one-day “Discovery Day in Health Sciences” event at the University of Calgary. Participants were randomly assigned to either the Verbal 6-Minute Walking Test (6MWT) or Music-Guided 6-minute walking (MU) cohorts. All SST and DTT instructions were delivered via the Ambulosono app operated on a smartphone to ensure uniformity in testing conditions with minimum human interference (Hu, 2019).

### Ambulosono Device

The Ambulosono device, a sensor-based kinematic measurement tool, utilizes high-precision accelerometers and gyroscopes to synchronize self-initiated movements with auditory instructions from an extensive acoustic library stored on the user’s mobile device [14]. Medically prescribed gait or joint rehabilitation protocols can be seamlessly integrated into the Ambulosono application, thereby enabling remote, home-based adherence to therapeutic regimens and ensuring consistency in both protocol administration and the testing environment (Chomiak et al., 2015). For this study, the Ambulosono system was employed to ensure standardized and consistent protocol delivery. Intervention-specific protocols were pre-loaded onto the device corresponding to the assigned intervention group.

### Participants

The study cohort comprised 54 high school students with a mean age of 15.98 years (SD = ±0.77 years). Participants were recruited via the Canadian Medical Hall of Fame and did not present with any physical or cognitive impairments that could potentially confound the SST or the intervention. Students were randomly allocated into one of two intervention arms: Verbal 6MWT (n=29) or MU (n=25).

### Procedure

Intervention-specific protocols were pre-loaded onto the Ambulosono application, and the Ambulosono sensor was affixed superior to either the left or right patella for the entire duration of the experimental protocol. Upon group allocation, each participant completed the SST on four separate occasions: pre-intervention (Captures 1 and 2) and post-intervention (Captures 3 and 4). Captures 1 and 3, denoted as single-task tests, consisted of a standard SST without stepping in place. Captures 2 and 4, designated as dual-task tests, required concurrent stepping-in-place and SST execution. After completing Captures 1 and 2, participants performed either the Verbal 6MWT or MU intervention, followed by Captures 3 and 4. 6MWT instructions were constructed according to the American Thoracic Society (ATS) guidelines (ATS, 2002; Chomiak et al., 2019). Participants in the Verbal 6MWT intervention group were first given general tests and safety instructions. During the test, they were notified about each minute they walked, together with pre-recorded words of encouragement, as well as the commands of altering gait speed during each minute from “walk at your comfortable speed” to “walk as fast as you can.” Two popular songs (“Yummy” by Justin Bieber and “Someone You Loved” by Lewis Capaldi) were used for the MU group.

### Data Collection and Analysis

During the SST iterations, the research team manually recorded and categorized participant responses into ‘Total Number of Answers’ and ‘Number of Incorrect Answers.’ All statistical computations were executed utilizing SPSS software for data analysis.

### Results Data Exclusion and Descriptive Statistics

A total of 11 data entries were deemed ineligible for analysis due to incompleteness, culminating in a final dataset comprising 43 valid entries (Verbal 6MWT: n=23; MU: n=20). Demographic information and descriptive statistics are delineated in Table 1-3.

**Table 1:** Demographic information of All High School Participants.

| Category | Group | Count | Percentage |
| --- | --- | --- | --- |
| Age (yrs) | 14 | 1 | 2.32% |
|  | 15 | 10 | 23.25% |
|  | 16 | 20 | 46.51% |
|  | 17 | 11 | 25.58% |
|  | 18 | 1 | 2.32% |
| Sex | Female | 29 | 67.44% |
|  | Male | 12 | 27.91% |
|  | Other | 2 | 4.64% |
| Height (cm) | 150-160 | 7 | 16.27% |
|  | 161-170 | 19 | 44.18% |
|  | 171-180 | 12 | 27.91% |
|  | 181-190 | 5 | 11.63% |
| Leg Length (cm) | 80 - 88 | 17 | 39.53% |
|  | 89 - 97 | 20 | 46.51% |
|  | 98 - 106 | 6 | 13.95% |

**Table 2:** Results from Wilcoxon Signed-Rank Test for the “Number of Total Answers” and “Number of Incorrect Answers”.

| Test Group | Comparison | Metric | p-value |
| --- | --- | --- | --- |
| Verbal 6MWT Group | Capture 1/Capture 3 | Total number of answers | 0.002* |
|  |  | Number of incorrect answers | 0.341 |
|  | Capture 2/Capture 4 | Total number of answers | 0.299 |
|  |  | Number of incorrect answers | 0.013* |
| MU Walking Test Group | Capture 1/Capture 3 | Total number of answers | 0.153 |
|  |  | Number of incorrect answers | 0.715 |
|  | Capture 2/Capture 4 | Total number of answers | 0.331 |
|  |  | Number of incorrect answers | 1.0 |

**Table 3:** Wilcoxon Signed-Rank Test Comparing the Percentage of Wrong Answers of Different Captures.

| Test Group | Comparison | P-Value |
| --- | --- | --- |
| Verbal 6MWT Group | Capture 1/Capture 3 | 0.826 |
|  | Capture 2/Capture 4 | 0.027* |
| MU Walking Group | Capture 1/Capture 3 | 0.650 |
|  | Capture 2/Capture 4 | 0.722 |

**Table 4:** Walking Data of the Interventions.

|  | MU Walking |  |  | 6MWT |  |  |
| --- | --- | --- | --- | --- | --- | --- |
|  | Walking Distance (km) | Walking Time (min) | Walking Speed (m/min) | Walking Distance (km) | Walking Time (min) | Walking Speed (m/min) |
| Average | 0.50 | 6.59 | 75.22 | 0.50 | 5.84 | 81.34 |
| Total | 9.42 | 125.13 | N/A | 10.01 | 134.26 | N/A |

### Normality Testing

The Shapiro-Wilk test was used to evaluate the null hypothesis that the dataset adheres to a normal distribution. Except for the total number of responses in the MU Walking Test Group (Capture 1/Capture 3 and Capture 2/Capture 4), all the other data variables are not normally distributed (P<0.05). As such the Wilcoxon signed-rank test was utilized to compare related samples or repeated measurements within individual samples such as the differences in scores between the pre- and post-intervention stages for each of the intervention groups (Verbal 6MWT and MU walking test) and each of the captures (Capture 1/Capture 3 and Capture 2/Capture 4).

### Standardizing Study Results

As participants provided varying numbers of both total answers and incorrect answers, those who gave fewer answers may have also provided fewer incorrect responses. Consequently, TC and XC standardized both the total number of answers and the number of wrong answers to the percentage of incorrect answers. Subsequently, XC conducted another set of Wilcoxon signed tests on the percentage of incorrect answers (Table 3). The comparisons between Capture 2 and 4 in the 6MWT revealed a statistically significant difference.

### Verbal 6MWT Group

Under single-task conditions (Capture 1/Capture 3), a statistically significant augmentation in the Total Number of Answers was observed post-intervention (p-value = 0.002), albeit without a corresponding change in the Number of Incorrect Answers (p-value = 0.341). However, the percentage of wrong answers did not significantly change in the post-intervention group (Figure 4). Under dual-task conditions (Capture 2/Capture 4), a significant alteration in the Number of Incorrect Answers was noted post-intervention (p-value = 0.013), without a significant change in the Total Number of Answers (p-value = 0.299). When comparing the percentage of wrong answers for both single-task and dual-task conditions, there is a statistically significant change in the dual-task conditions, indicating improvement post-intervention (Figure 5).

### MU Group

Under single-task conditions (Capture 1/Capture 3), no statistically significant differences were observed in either the Total Number of Answers (p-value = 0.153) or the Number of Incorrect Answers (p-value = 0.715) post-intervention. Under dual-task conditions (Capture 2/Capture 4), the intervention did not yield any statistically significant differences in either the Total Number of Answers (p-value = 0.331) or the Number of Incorrect Answers (p-value = 1.0). Under both conditions, the intervention yielded no statistically significant difference in the percentage of wrong answers (Figures 2 and 3).

**FIGURE 1:**
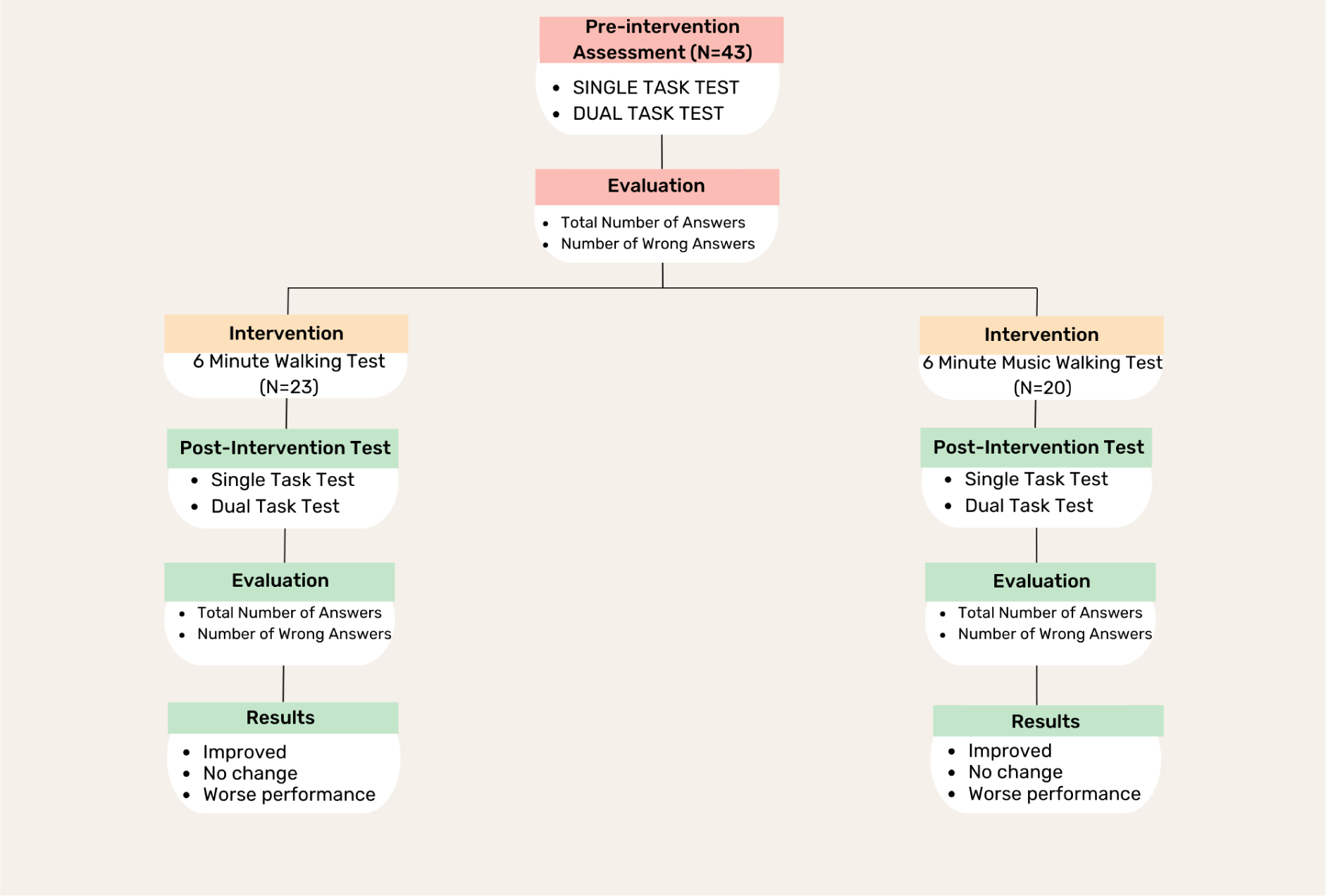
DATA COLLECTION PROCESS.

**Figure 2:**
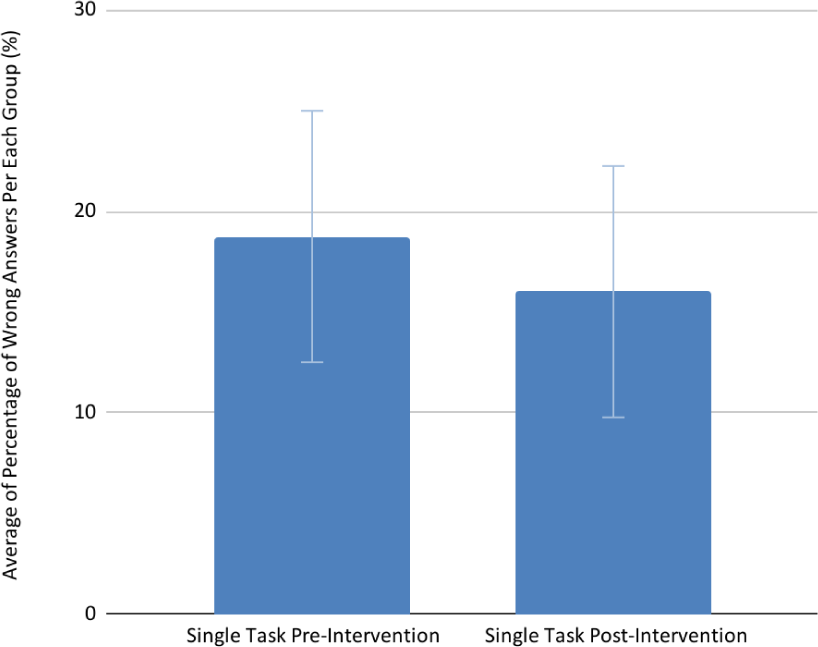
Comparison of the Average of Percentage of Wrong Answers Between Single Task Before and After Music Walking (CP1 vs CP3)

**Figure 3:**
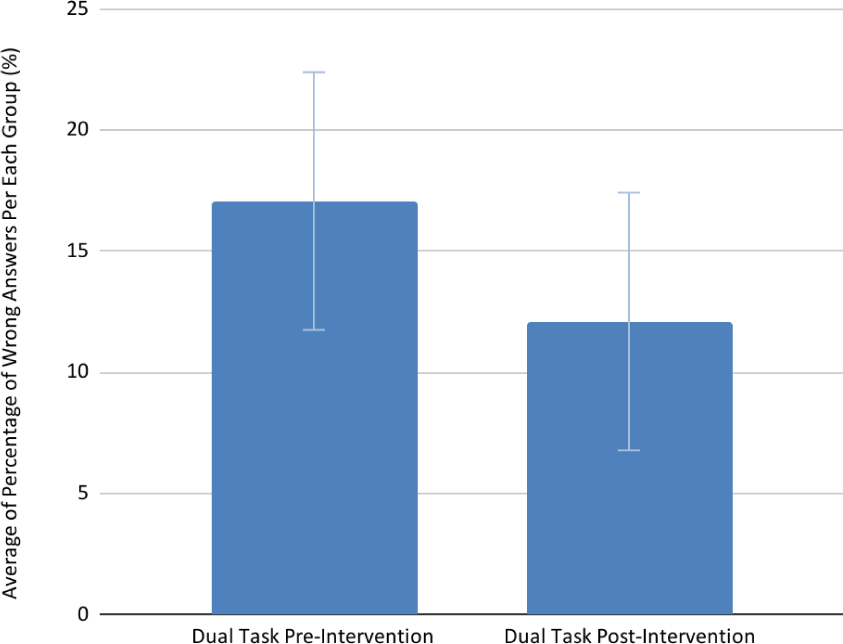
Comparison of the Average of Percentage of Wrong Answers Between Dual Task Before and After Music Walking (CP2 vs Cp4)

**Figure 4:**
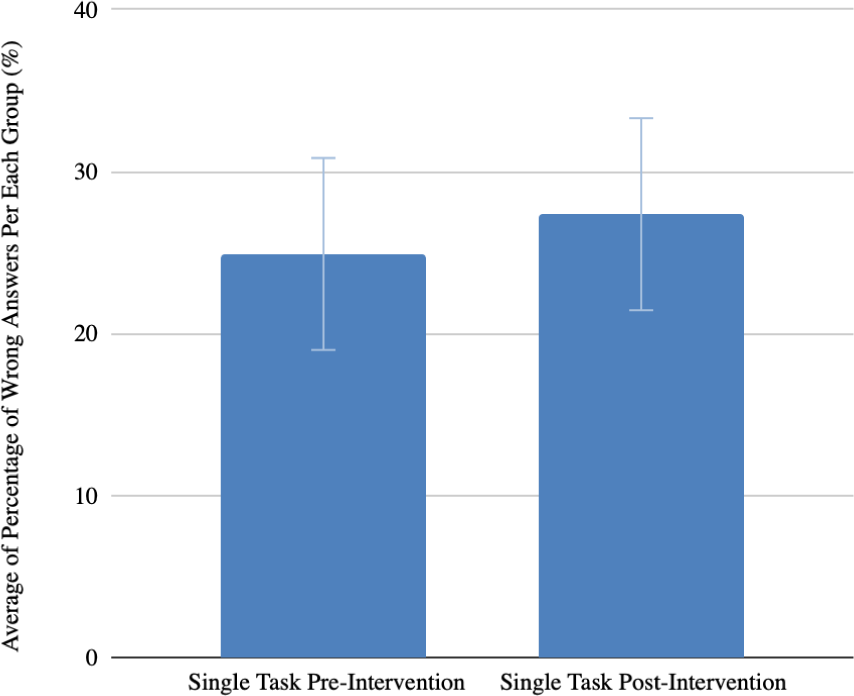
Comparison of the Average of Percentage of Wrong Answers Between Single Taks Before and After 6 Minutes Walking Test (CP1 vs CP3)

**Figure 5:**
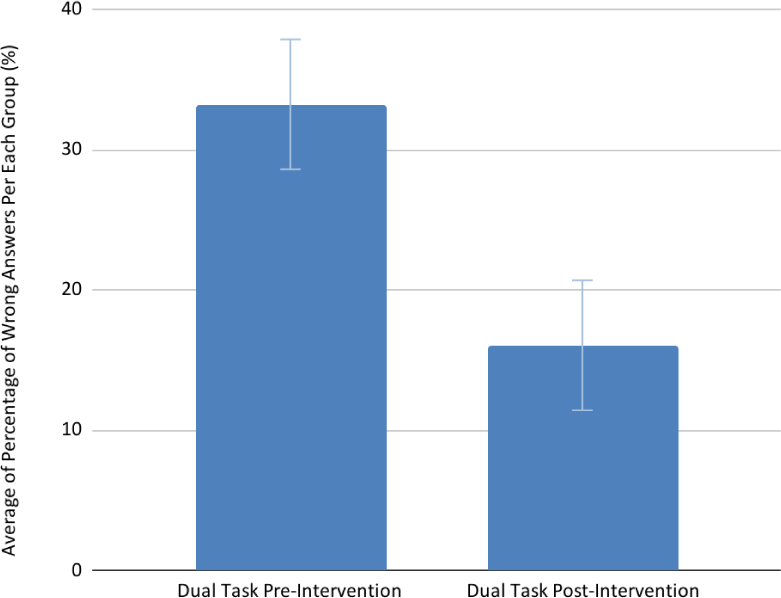
Comparison of the Average of Percentage of Wrong Answers Between Dual Task Before and After 6 Minutes Walking Test (CP2 vs CP4)

### Walking Data Comparisons

To ensure similar exercise intensity for the two intervention groups, we examined the difference between the walking data (walking distance, time, and speed) obtained by the Ambulosono sensors. The Mann-Whitney test did not yield any significant differences for the two intervention groups. Spearman’s Nonparametric Correlation tests were also performed to explore correlations between walking data and participants’ performance on the SST, which yielded no significant results.

## Discussion

### Overview and Methodological Approach

The Serial Subtract Test (SST) is a gold standard tool for appraising cognitive performance in varied research and clinical contexts. Utilizing the Ambulosono device and app-controlled verbal speed instructions, our study was able to standardize walking speed among subjects, a key factor that may influence exercise intensity if subjects choose to walk at different speeds.

### Differential Effects of Physical Activity and Music on Cognitive Performance

In a previous study, Stewart (2006) discussed the intricate relationship between physical activity, cognition, and music. Our study is largely consistent with this view: physical activity associated with 6MWT can lead to enhanced cognitive performance under both single and dual-task conditions. This positive exercise effect was however absent in the MU intervention group. This phenomenon cannot be ascribed to the distance nor the speed of walking because both groups have similar exercise intensities based on movement data captured by Ambulosono sensors.

Despite abundant evidence supporting music’s effect on cognitive capabilities (Chang et al., 2012; Heyn et al., 2004; Verrusio et al., 2015), our results highlight the complexities of music on cognitive performance. Music listening can activate brain areas associated with memory and attention, which are similarly engaged during the SST (Chomiak et al., 2015; Karzmark, 2000; Verrusio et al., 2015). Consequently, the MU group may have experienced a higher cognitive load compared to 6MWT due to listening to music, which could potentially divert cognitive resources away from the SST and DTT capabilities (Balogun et al., 2013; Steward et al., 2006) and impact their post-intervention performance. Another possible reason could be that students in the MU walking group had better mathematical ability compared to the 6MWT group as the MU walking group started with a higher baseline performance (Watts et al., 2014).

Conversely, the improved performance in single-task and dual-task conditions observed in the 6MWT group could be attributed to a lower cognitive load compared to the MU intervention group. This aligns with studies, such as by Mualem et al. (2018), highlighting brief walking sessions’ cognitive benefits across demographics (Mualem et al., 2018). Indeed, several meta-analyses further indicate that exercise can produce positive cognitive benefits (Chang et al., 2012; Colcombe & Kramer, 2003; Heyn et al., 2004; Xu et al., 2023). Another possible explanation is that the MU walking group began with a lower percentage of wrong answers during Capture 1, the initial test, compared to the 6MWT group, suggesting a ceiling effect (Penko et al., 2021).

### Differential Effects of Exercise on Single vs. Dual-Task Performance

We found that in the single-task conditions (Capture 1/Capture 3), the 6MWT exercise group increased the Number of Total Answers but had no effect on the Number of Incorrect Answers. In contrast, in the dual-task condition (Capture 2/Capture 4), the intervention did not significantly change the Number of Total Answers but significantly reduced the number of wrong answers given by the participants. The divergent outcomes between the single task and dual task conditions warrant further examination. It is possible that such a differential effect may simply reflect the fact that different aspects of SST are differentially attended under single and dual-task conditions, with the correctness of the answers being more attended by the subjects under the dual-tasking condition (Aliakbaryhosseinabadi et al., 2017).

### Implications

To our knowledge, our study is the first that systematically explored the potential of wearable devices and apps in studying exercise and cognition. The Ambulosono technology not only allowed us to develop protocols to integrate cognitive tests and exercise intervention but also controls for exercise intensity and emotional interference. The versatility and quantitative nature of the Ambulosono digital tool not only substantially enhances the quality of research but can be further explored in real-world patient care. Indeed, the findings that brief exercise of 6MWT and the improved cognitive performance suggest the intervention could help patients grappling with cognitive or mobility challenges, such as Parkinson’s or Alzheimer’s disease (Xu et al., 2023).

### Study Limitations

While our research presents fresh insights, the limited sample size might diminish its statistical robustness, such as the muted outcomes in the MU walking group. Furthermore, a study with large sample size and different forms of exercise is needed to validate the cognitive benefits in SST and DTT and for more intense exercise than 6MWT.

### Conclusion

Our findings indicate that the digital technology-led verbal 6-Minute Walking Test (6MWT) intervention outperforms the MU intervention in enhancing cognitive performance. Specifically, the 6MWT significantly improved the total number of correct answers for the single-task condition post-intervention and reduced the number of incorrect answers for the dual-task condition post-intervention, whereas the MU intervention did not yield significant changes in either metric under single- or dual-task conditions. Our study presents a comprehensive exploration into the intricate relationship between physical activity, cognitive function, and external stimuli such as music, using the Serial Subtraction Test (SST) as a benchmark for cognitive performance. Utilizing the Ambulosono device’s digital health technology, we standardized brisk walking exercises under varying conditions, including single-task and dual-task scenarios with and without musical accompaniment. This research not only corroborates the cognitive benefits of physical activity but also introduces nuanced insights into the role of external stimuli like music. It paves the way for future investigations to optimize cognitive interventions, thereby contributing to the broader scientific discourse on the nexus between physical activity, cognitive function, and digital health technology.

## Data Availability

All data produced in the present study are available upon reasonable request to the authors

## Acknowledgments

We extend our sincere gratitude to Adam David for his invaluable advice and guidance throughout the research process, as well as for his pivotal role in organizing the research collection event. Our thanks also go to Janice Morgan for her essential contribution in connecting us with high school students and coordinating the Discovery Day event, which was crucial for our data collection. We would like to express our appreciation to the Scholars Academy Program at the University of Calgary, and specifically to Howard, Gala, Rose, Marcela, Laurine, and Amber, for their assistance and support in data gathering during the Discovery Day event. Lastly, our heartfelt thanks to all the high school students who participated in our study. Your involvement was fundamental to the success of our research.

## Notes

### Competing Interest Statement

The authors have declared no competing interest.

### Clinical Trial

ISRCTN06023392

### Funding Statement

This study was funded by Alberta Ministry of Mental Health

### Author Declarations

Ethics committee/IRB of University of Calgary waived ethical approval for this work

## References

Åhman, H. B., Cedervall, Y., Kilander, L., Giedraitis, V., Berglund, L., McKee, K. J., Rosendahl, E., Ingelsson, M., & Åberg, A. C. (2020). Dual-task tests discriminate between dementia, mild cognitive impairment, subjective cognitive impairment, and healthy controls – a cross-sectional cohort study. BMC Geriatrics, 20(1). 10.1186/s12877-020-01645-1

Åhman, H. B., Giedraitis, V., Cedervall, Y., Lennhed, B., Berglund, L., McKee, K., Kilander, L., Rosendahl, E., Ingelsson, M., & Åberg, A. C. (2019). Dual-task performance and neurodegeneration: Correlations between timed up-and-go dual-task test outcomes and alzheimer’s disease cerebrospinal fluid biomarkers. Journal of Alzheimer’s Disease, 71(s1). 10.3233/jad-181265

Aliakbaryhosseinabadi, S., Kamavuako, E. N., Jiang, N., Farina, D., & Mrachacz-Kersting, N. (2017). Influence of dual-tasking with different levels of attention diversion on characteristics of the movement-related cortical potential. Brain Research, 1674, 10–19. 10.1016/j.brainres.2017.08.016

Al-Yahya, E., Dawes, H., Smith, L., Dennis, A., Howells, K., & Cockburn, J. (2011). Cognitive motor interference while walking: A systematic review and meta-analysis. Neuroscience & Biobehavioral Reviews, 35(3), 715–728. 10.1016/j.neubiorev.2010.08.008

Alter, D. A., O’Sullivan, M., Oh, P. I., Redelmeier, D. A., Marzolini, S., Liu, R., Forhan, M., Silver, M., Goodman, J. M., & Bartel, L. R. (2015). Synchronized personalized music audio-playlists to improve adherence to physical activity among patients participating in a structured exercise program: A proof-of-principle feasibility study. Sports Medicine - Open, 1(1). 10.1186/s40798-015-0017-9

American Thoracic Society Committee on Proficiency Standards for Clinical Pulmonary Function Laboratories. (2002). ATS statement: Guidelines for the six-minute walk test. American Journal of Respiratory and Critical Care Medicine, 166(1), 111–117. 10.1164/ajrccm.166.1.at1102

Angel, L. A., Polzella, D. J., & Elvers, G. C. (2010). Background music and cognitive performance. Perceptual and Motor Skills, 110(3), 1059–1064. 10.2466/pms.110.c.1059-1064

Ballmann, C. G. (2021). The influence of Music Preference on exercise responses and performance: A Review. Journal of Functional Morphology and Kinesiology, 6(2), 33. 10.3390/jfmk6020033

Balogun S. K., Monteiro N. M., Tseletso T. (2013). Effects of music genre and music language on task performance among University of Botswana Students. American Journal of Applied Psychology, 1(3), 38–43. https://ezproxy.usc.edu.au//dx.doi.org/10.12691/ajap-1-3-2

Chang, Y. K., Labban, J. D., Gapin, J. I., & Etnier, J. L. (2012). The effects of acute exercise on cognitive performance: A meta-analysis. Brain Research, 1453, 87–101. 10.1016/j.brainres.2012.02.068

Chang, Y.-K., & Etnier, J. L. (2009). Exploring the dose-response relationship between resistance exercise intensity and cognitive function. Journal of Sport and Exercise Psychology, 31(5), 640–656. 10.1123/jsep.31.5.640

Chomiak, T., Pereira, F. V., Meyer, N., de Bruin, N., Derwent, L., Luan, K., Cihal, A., Brown, L. A., & Hu, B. (2015). A new quantitative method for evaluating freezing of gait and dual-attention task deficits in parkinson’s disease. Journal of Neural Transmission, 122(11), 1523–1531. 10.1007/s00702-015-1423-3

Chomiak, T., Sidhu, A., Watts, A., Su, L., Graham, B., Wu, J., Classen, S., Falter, B., & Hu, B. (2019). Development and validation of Ambulosono: A wearable sensor for bio-feedback rehabilitation training. Sensors, 19(3), 686. 10.3390/s19030686

Colcombe, S. J., Erickson, K. I., Scalf, P. E., Kim, J. S., Prakash, R., McAuley, E., Elavsky, S., Marquez, D. X., Hu, L., & Kramer, A. F. (2006). Aerobic exercise training increases brain volume in aging humans. The Journals of Gerontology Series A: Biological Sciences and Medical Sciences, 61(11), 1166–1170. 10.1093/gerona/61.11.1166

Colcombe, S., & Kramer, A. F. (2003). Fitness effects on the cognitive function of older adults. Psychological Science, 14(2), 125–130. 10.1111/1467-9280.t01-1-01430

Curzel, J., Forgiarini Junior, L. A., & Rieder, M. de. (2013). Evaluation of functional independence after discharge from the Intensive Care Unit. Revista Brasileira de Terapia Intensiva, 25(2), 93–98. 10.5935/0103-507x.20130019

Erickson, K. I., Voss, M. W., Prakash, R. S., Basak, C., Szabo, A., Chaddock, L., Kim, J. S., Heo, S., Alves, H., White, S. M., Wojcicki, T. R., Mailey, E., Vieira, V. J., Martin, S. A., Pence, B. D., Woods, J. A., McAuley, E., & Kramer, A. F. (2011). Exercise training increases size of hippocampus and improves memory. Proceedings of the National Academy of Sciences, 108(7), 3017–3022. 10.1073/pnas.1015950108

Graham, J. D., Li, Y.-C., Bray, S. R., & Cairney, J. (2018). Effects of cognitive control exertion and motor coordination on task self-efficacy and muscular endurance performance in children. Frontiers in Human Neuroscience, 12. 10.3389/fnhum.2018.00379

Hayman, M. (1942). Two minute clinical test for measurement of intellectual impairment in psychiatric disorders. Archives of Neurology And Psychiatry, 47(3), 454. 10.1001/archneurpsyc.1942.02290030112010

Herold, F., Törpel, A., Hamacher, D., Budde, H., Zou, L., Strobach, T., Müller, N. G., & Gronwald, T. (2021). Causes and consequences of interindividual response variability: A call to apply a more rigorous research design in acute exercise-cognition studies. Frontiers in Physiology, 12. 10.3389/fphys.2021.682891

Heyn, P., Abreu, B. C., & Ottenbacher, K. J. (2004). The effects of exercise training on elderly persons with cognitive impairment and dementia: A meta-analysis. Archives of Physical Medicine and Rehabilitation, 85(10), 1694–1704. 10.1016/j.apmr.2004.03.019

Hu, B. (2019). Application of wearable technology in clinical walking and dual task testing. Journal of Translational Internal Medicine, 7(3), 87–89. 10.2478/jtim-2019-0019

Hu, B., & Chomiak, T. (2019). Wearable technological platform for multidomain diagnostic and exercise interventions in parkinson’s disease. International Review of Neurobiology, 75–93. 10.1016/bs.irn.2019.08.004

Karzmark, P. (2000). Validity of the serial seven procedure. International Journal of Geriatric Psychiatry, 15(8), 677–679. 10.1002/1099-1166(200008)15:8<677::aid-gps177>3.0.co;2-4

Keye, S. A., Walk, A. M., Cannavale, C. N., Iwinski, S., McLoughlin, G. M., Steinberg, L. G., & Khan, N. A. (2021). Six-minute walking test performance relates to neurocognitive abilities in preschoolers. Journal of Clinical Medicine, 10(4), 584. 10.3390/jcm10040584

Mammarella, N., Fairfield, B., & Cornoldi, C. (2007). Does music enhance cognitive performance in healthy older adults? the Vivaldi effect. Aging Clinical and Experimental Research, 19(5), 394–399.10.1007/bf03324720

Mandolesi, L., Polverino, A., Montuori, S., Foti, F., Ferraioli, G., Sorrentino, P., & Sorrentino, G. (2018). Effects of physical exercise on cognitive functioning and wellbeing: Biological and psychological benefits. Frontiers in Psychology, 9. 10.3389/fpsyg.2018.00509

Montero-Odasso, M., Zou, G., Speechley, M., Almeida, Q. J., Liu-Ambrose, T., Middleton, L. E., Camicioli, R., Bray, N. W., Li, K. Z. H., Fraser, S., Pieruccini-Faria, F., Berryman, N., Lussier, M., Shoemaker, J. K., Son, S., Bherer, L., McFadyen, B. J., Barha, C., & McGibbon, C. (2023). Effects of exercise alone or combined with cognitive training and vitamin D supplementation to improve cognition in adults with mild cognitive impairment. JAMA Network Open, 6(7). 10.1001/jamanetworkopen.2023.24465

Mualem, R., Leisman, G., Zbedat, Y., Ganem, S., Mualem, O., Amaria, M., Kozle, A., Khayat-Moughrabi, S., & Ornai, A. (2018). The effect of movement on cognitive performance. Frontiers in Public Health, 6.10.3389/fpubh.2018.00100

Mualem, R., Leisman, G., Zbedat, Y., Ganem, S., Mualem, O., Amaria, M., Kozle, A., Khayat-Moughrabi, S., & Ornai, A. (2018). The effect of movement on cognitive performance. Frontiers in Public Health, 6, 100–100. 10.3389/fpubh.2018.00100

Murman, D. (2015). The impact of age on cognition. Seminars in Hearing, 36(03), 111–121. 10.1055/s-0035-1555115

Plummer, P., & Eskes, G. (2015). Measuring treatment effects on dual-task performance: A framework for research and clinical practice. Frontiers in Human Neuroscience, 9. 10.3389/fnhum.2015.00225

Roy, E. (2013). Cognitive function. Encyclopedia of Behavioral Medicine, 448–449. 10.1007/978-1-4419-1005-9_1117

Särkämö, T. (2018). Cognitive, emotional, and neural benefits of musical leisure activities in aging and neurological rehabilitation: A critical review. Annals of Physical and Rehabilitation Medicine, 61(6), 414–418. 10.1016/j.rehab.2017.03.006

Sperandio, E. F., Arantes, R. L., Matheus, A. C., Silva, R. P., Lauria, V. T., Romiti, M., Gagliardi, A. R. T., & Dourado, V. Z. (2015). Intensity and physiological responses to the 6-minute walk test in middle-aged and older adults: A comparison with cardiopulmonary exercise testing. Brazilian Journal of Medical and Biological Research, 48(4), 349–353. 10.1590/1414-431x20144235

Srygley, J. M., Mirelman, A., Herman, T., Giladi, N., & Hausdorff, J. M. (2009). When does walking alter thinking? age and task associated findings. Brain Research, 1253, 92–99. 10.1016/j.brainres.2008.11.067

Stewart, L., von Kriegstein, K., Warren, J. D., & Griffiths, T. D. (2006). Music and the brain: Disorders of musical listening. Brain, 129(10), 2533–2553. 10.1093/brain/awl171

Thompson, R. G., Moulin, C. J., Hayre, S., & Jones, R. W. (2005). Music enhances category fluency in healthy older adults and alzheimer’s disease patients. Experimental Aging Research, 31(1), 91–99. 10.1080/03610730590882819

Verrusio, W., Ettorre, E., Vicenzini, E., Vanacore, N., Cacciafesta, M., & Mecarelli, O. (2015). The mozart effect: A quantitative EEG study. Consciousness and Cognition, 35, 150–155. 10.1016/j.concog.2015.05.005

Voelcker-Rehage, C., Niemann, C., Hübner, L., Godde, B., & Winneke, A. H. (2016). Benefits of physical activity and fitness for lifelong cognitive and motor development—brain and behavior. Sport and Exercise Psychology Research, 43–73. 10.1016/b978-0-12-803634-1.00003-0

Wang, R., Zhang, H., Li, H., Ren, H., Sun, T., Xu, L., Liu, Y., & Hou, X. (2022). The influence of exercise interventions on cognitive functions in patients with amnestic mild cognitive impairment: A systematic review and meta-analysis. Frontiers in Public Health, 10. 10.3389/fpubh.2022.1046841

Watts, T. W., Duncan, G. J., Siegler, R. S., & Davis-Kean, P. E. (2014). What’s past is prologue. Educational Researcher, 43(7), 352–360. 10.3102/0013189x14553660

Wu, G., Sanderson, B., & Bittner, V. (2003). The 6-minute walk test: How important is the learning effect? American Heart Journal, 146(1), 129–133. 10.1016/s0002-8703(03)00119-4

Xu, L., Gu, H., Cai, X., Zhang, Y., Hou, X., Yu, J., & Sun, T. (2023). The effects of exercise for cognitive function in older adults: A systematic review and meta-analysis of randomized controlled trials. International Journal of Environmental Research and Public Health, 20(2), 1088. 10.3390/ijerph20021088

Yang, L., He, C., & Pang, M. Y. (2016). Reliability and validity of dual-task mobility assessments in people with chronic stroke. PLOS ONE, 11(1). 10.1371/journal.pone.0147833

Yogev-Seligmann, G., Hausdorff, J. M., & Giladi, N. (2007). The role of executive function and attention in gait. Movement Disorders, 23(3), 329–342. 10.1002/mds.21720

